# Genetic and functional evidence implicates a *CRIP3* non-synonymous variant in age-related hearing loss

**DOI:** 10.64898/2025.12.22.25342816

**Authors:** Sydney E. Brongo, Alber Aqil, Matthew A. Xu-Friedman, Omer Gokcumen

**Affiliations:** Department of Biological Sciences, State University of New York at Buffalo, Buffalo, NY 14260; Department of Computational Medicine and Bioinformatics, University of Michigan, Ann Arbor, MI, 48109

**Keywords:** Age-related hearing loss, *CRIP3*, Functional genomics, GWAS, Antagonistic pleiotropy

## Abstract

Age-related hearing loss is a widespread sensory impairment affecting a significant proportion of the elderly population, yet the genetic underpinnings of this condition remain incompletely understood. In this study, we investigate a non-synonymous variant (rs2242416) in the *CRIP3* gene, which is expressed in auditory hair cells, in the context of hearing loss. Firstly, we find the variant shows strong and consistent association with hearing loss across multiple genome-wide association studies. Secondly, this variant, substitutes the nonpolar isoleucine for the polar threonine at an amino acid site that is otherwise highly conserved across placental mammals. Thirdly, by causing the amino acid change, the variant subtly alters the structure of the CRIP3 protein. Together, these three analyses provide phenotypic, evolutionary, and molecular evidence for the functionality of *CRIP3* and its role in hearing loss. Moreover, the population genetics of the *CRIP3* locus reveals an increased frequency of the derived threonine allele of rs2242416 in Eurasian populations following the out-of-Africa migration of humans more than 50,000 years ago. Nevertheless, the role of Darwinian selection in this increased frequency remains inconclusive. Overall, our results make a compelling argument to auditory researchers to make a *CRIP3* mouse model to pinpoint the precise role of the protein in auditory function. Such a model will pave the way for therapeutic interventions targeting the CRIP3 protein to mitigate age-related hearing loss.

## Introduction

Age-related hearing loss is one of the most prevalent forms of sensory impairment, affecting approximately one in three adults over the age of 65 [1]. In addition to being a sensory deficit, hearing loss has been strongly linked to increased risks of social isolation and cognitive decline [2, 3]. These represent enormous costs to society, so it is important to identify causes of age-related hearing loss so that therapies and prevention strategies can be developed.

A variety of factors contribute to hearing loss, including environmental exposures such as noise, ototoxic medications, and genetic predisposition [4]. Many proteins critical to hearing have been identified through their roles in non-syndromic deafness [5, 6]. Recent genome-wide association studies (GWAS) have provided valuable insights into the complex genetic architecture of hearing loss [7-9]. Many genes have been associated with age-related hearing loss, with genetic estimates ranging from as low as 30% to as high as 70% [8, 10-16]. This inconsistency stresses the need to identify further genetic risks.

In this study, we identified a common haplotype that harbors variants that are strongly associated with an increased risk of hearing loss [7-9], including a non-synonymous mutation in the *CRIP3* gene, which encodes cysteine-rich protein 3. CRIP3 is a LIM-domain protein that was first described in 2001. While its function is still unclear, members of the CRIP family are known to mediate protein-protein interactions and association with actin filaments [17-20]. Our evolutionary analyses suggest that the high frequency of this variant in human populations may be explained, at least in part, by historical selective pressures. Further, we found evidence that *CRIP3* is expressed in both cochlear hair cells [9] and non-auditory structures, such as the left ventricle of the heart [21], suggesting potential pleiotropic functions.

Overall, the combination of functional genomics and evolutionary analyses suggest a key variant that contributes to age-related hearing loss and urges experimental follow-up with mouse models.

## Results

### *CRIP3* locus is associated with hearing loss and heart disease

To explore the genetic basis of hearing loss, we examined top genome-wide association study (GWAS) signals linked to hearing loss from the GWAS Atlas (**Figure S1**). Among these, two haplotypes overlapped protein-coding regions: one within *ARHGEF28* on chromosome 5 and another within *CRIP3/ZNF318* on chromosome 6. Inspection of trait-associated single-nucleotide variants (SNVs) in these loci highlighted rs2242416 as a strong candidate, as it lies within the *CRIP3* coding sequence and causes a rare amino acid change from nonpolar isoleucine (encoded by an ancestral “A”) to polar threonine (encoded by a derived “G”) [7-9].Such polarity-altering substitutions are uncommon among GWAS-identified variants and often have significant functional effects [22-24].

To validate the association between rs2242416 and hearing loss, we conducted a phenotype-wide association study (PheWAS). We observed that rs2242416 was strongly associated with hearing loss in both the GWAS Atlas (p = 3.69 × 10^−18^; **Table S1, Figure 1A**) and FinnGen ([25]; p = 4.53 × 10^−17^; **Figure S2**), specifically the derived “G” allele. Remarkably, the same variant was also associated with cardiovascular traits, including hypertension in FinnGen (p = 7.44 × 10^−13^) and vascular or heart problems in the GWAS Atlas (p = 2.20 × 10^−12^) (**Figure S2**), specifically the ancestral “A” allele. In other words, the allele that elevates hearing-loss risk shows the opposite effect on vascular traits.

**Figure 1.**
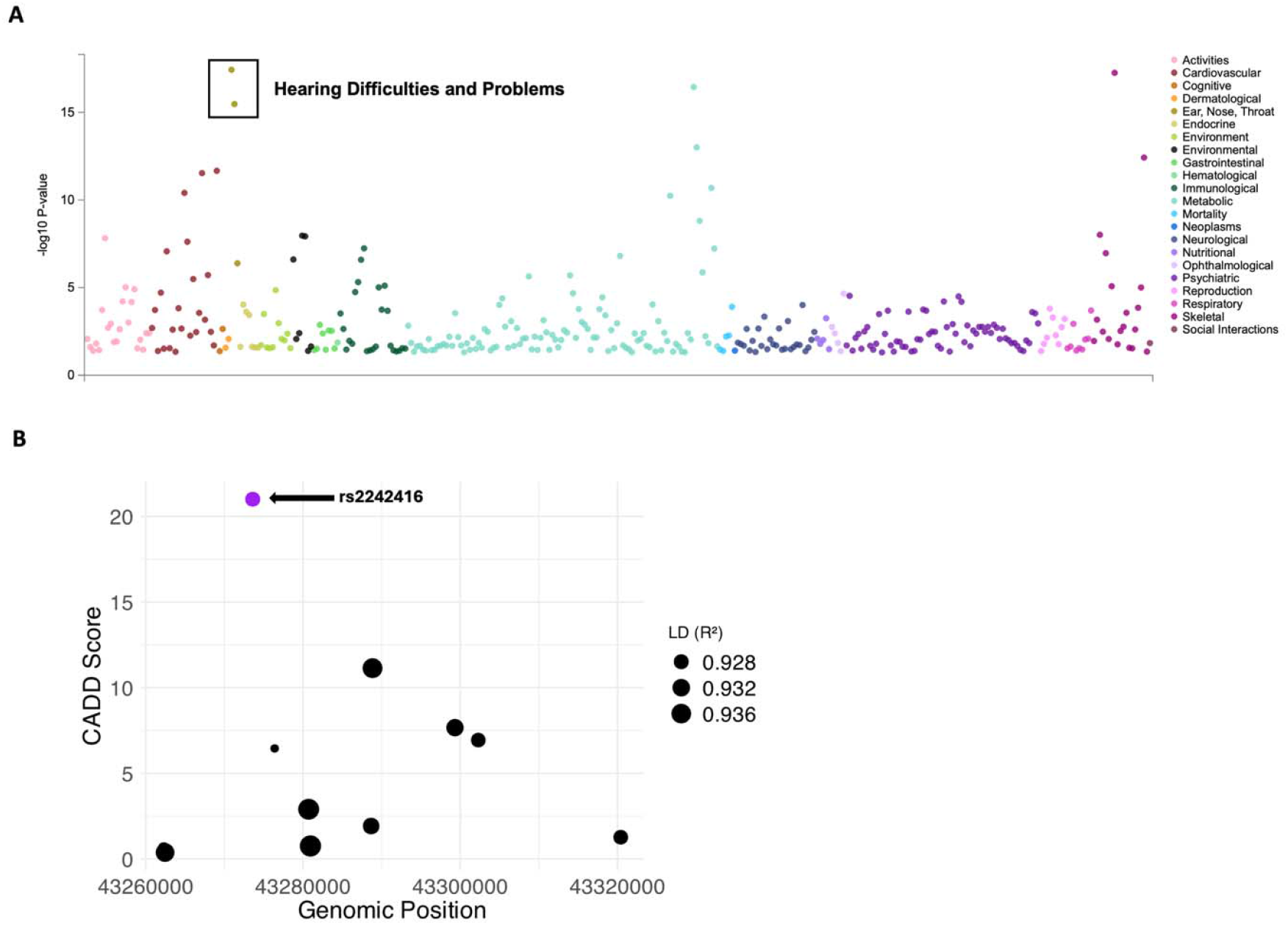
Confirmation of rs2242416 being a causal variant. A. PheWas plot showing top associations with rs2242416, with 2 out of 4 of the top associations being hearing difficulties and problems. B. Linkage disequilibrium scores (*r*^2^> 0.9) for variants in linkage disequilibrium with our variant of interest (rs2242416) and their CADD score. rs2242416 is highlighted in purple. A CADD score of 20 or greater denotes a substitution that is in the top 1% deleterious substitutions in the human genome.

In parallel, using the Genotype Tissue Expression (GTEx) database [21], we found that *CRIP3* is expressed in both the brain and heart, consistent with the associations for hearing and heart function. Furthermore, *CRIP3* expression has been detected specifically in auditory hair cells [9],further underscoring its potential role in auditory physiology. Further inquiry into the GTEX database, showed that the haplotype harboring rs2242416 is correlated with a significant decrease in expression of *CRIP3* in multiple tissues (p <10^−9^, **Figure S3B**). This result is consistent with previous findings that dramatic changes in expression levels driven by genetic variants can lead to disease states [26].

### In silico functional analysis identifies rs2242416 as the likely causal variant underlying the GWAS association

GWAS signals often reflect recombination blocks inherited together, making it crucial to identify the specific variant among linked alleles that drives the phenotypic association. To determine whether rs2242416 is the causal variant, that is, the specific DNA change directly responsible for altering *CRIP3* function and increasing susceptibility to hearing loss, we evaluated other genetic variants within the *CRIP3* haplotype. Variants that are physically close together on the chromosome are often inherited as a block resulting in the co-occurrence of those alleles across people, a phenomenon known as linkage disequilibrium (LD). Thus, when variants are tightly linked, it can be difficult to determine which variant is driving the biological effect. To address this, we identified ten variants in strong LD (*r*^2^> 0.9) with rs2242416 (**Figure 1B**). However, none of these variants showed comparable functional significance based on their Combined Annotation Dependent Deletion (CADD) scores, which estimates how damaging a genetic variant may be. These results support rs2242416 as the most likely causal variant for age-related hearing loss within the haplotype [27].

Next, we examined how this amino acid substitution might alter CRIP3 protein function. Using AlphaFold3 [28],we compared predicted CRIP3 models carrying the ancestral “A” allele (isoleucine) versus the derived “G” allele (threonine) (**Figure 2A**). The substitution is predicted to introduce an additional hydrogen bond when threonine replaces isoleucine, which may increase local structural stability [29].Although the precise function of CRIP3 remains unclear, this additional hydrogen bond could influence protein–protein interactions or stability.

**Figure 2.**
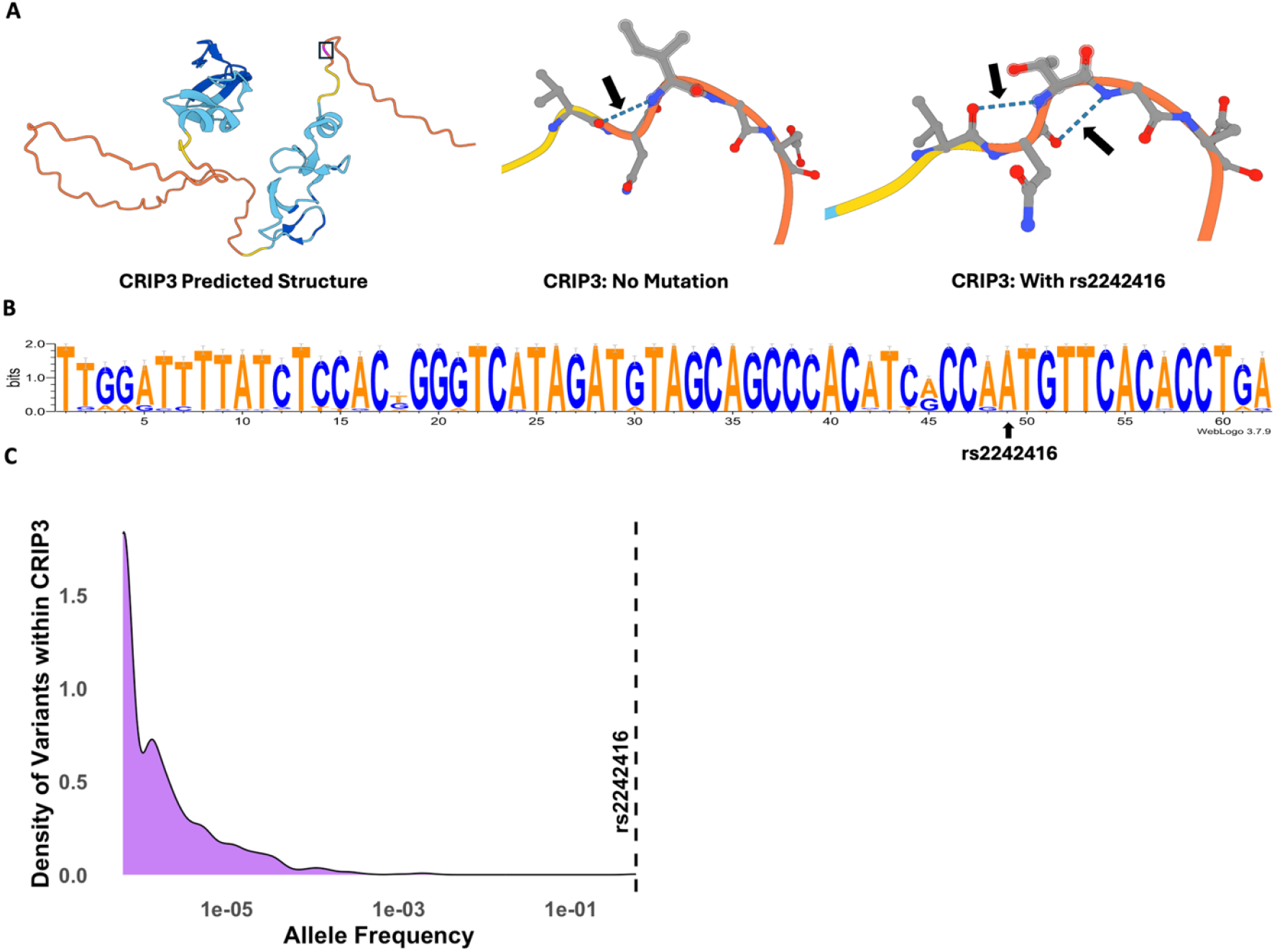
Structural Prediction and Distribution of rs2242416 across species. A. From left to right, CRIP3 predicted structure using AlphaFold3, Position 188 with 1 hydrogen bond (CRIP3: no mutation), Position 188 with 2 hydrogen bonds (CRIP3: with rs2242416). Colors of structures indicate reliability of structure prediction with blue being very high (predicted Local Distance Difference Test >90 (pLDDT)) and orange being very low (pLDDT< 50). B. Sequence logo showing the conservation of the A allele across multiple species. C. Distribution of variants within *CRIP3* in human populations and their corresponding allele frequencies. Notably, rs2242416 has the highest allele frequency of variants found within *CRIP3*.

Because protein sequences are typically conserved across species, non-synonymous substitutions that alter amino acid polarity, such as the isoleucine-to-threonine change caused by rs2242416, are particularly rare [30].To assess the evolutionary conservation of this site, we queried the CACTUS alignment database [31],which compiles multi-species conservation data across placental mammals. All examined species, except two lemurs and the rabbit (see Discussion), carry the ancestral “A” allele encoding isoleucine (**Figure 2B**). Further, other derived non-synonymous variants found in *CRIP3* are all extremely rare (<0.24%, **Figure 2C**). In contrast, the derived “G” allele encoding threonine occurs at more than 50% frequency across human populations in Eurasia (**Figure 3A**). This striking divergence from an otherwise conserved ancestral state may be driven by positive selection. We test this hypothesis in the following section.

**Figure 3.**
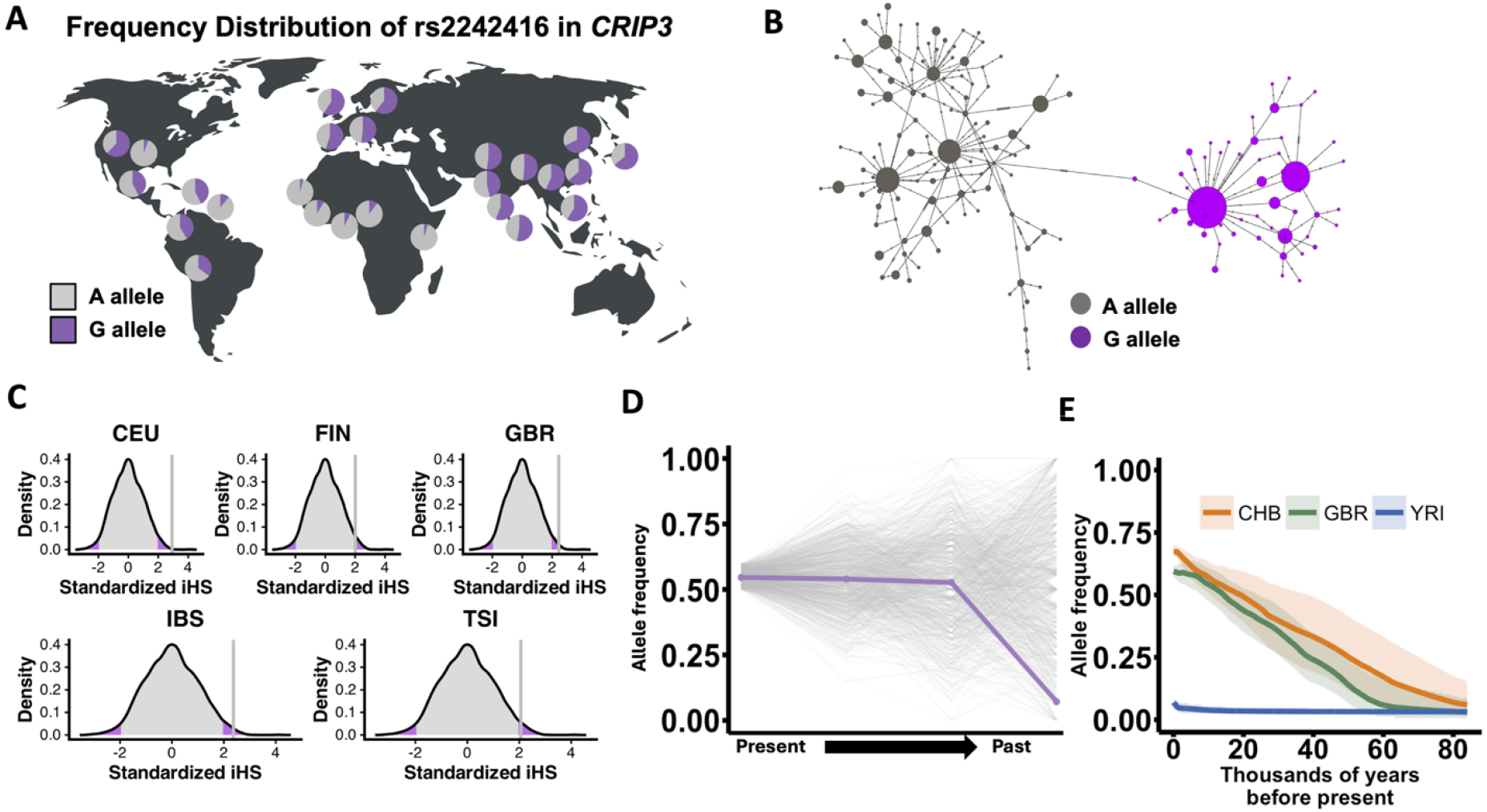
Signatures of Selections in European Populations. A. Frequency distribution of rs2242416 in multiple populations from the 1000 Genomes Project. B. Haplotype network of populations from the 1000 Genomes Project, both with and without rs2242416. Populations include Europeans (CEU: Utah residents with Northern and Western European ancestry, FIN: Finnish in Finland, TSI: Toscani in Italia, GBR: British in England and Scotland), East Asians (CHB: Han Chinese in Beijing, China, CHS: Han Chinese South,CDX: Denver Chinese, KHV: Kinh Vietnamese, JPT: Japanese), Africans (YRI: Yoruba, GWD: Gambian, MSL: Mende, ESN: Esan, LWK: Luhya) and South Asians(BEB: Bengali, PJL: Punjabi, ITU: Indian, STU: Sri Lankan, GIH: Gujarati). C. Standardized IHS values for European populations. Population codes: Refer to B and IBS: Iberian populations in Spain. The significance zones are shown in purple, and the Z-score for rs2242416 is shown as a vertical line on each graph. D. Allele frequency trajectory for the derived “G” allele. E. Clues analysis for 3 populations with log likelihood values and selection coefficients: CHB: *logLR*: 1.7206; s: 0.00110, GBR: *logLR*: 1.9249; *s*: 0.00094, YRI: *logLR*: 0.7077; *s*: 0.00498.

### Evolution of the rs2242416 variant

To test the hypothesis of positive selection at rs2242416, we first investigated the allele frequency distribution of this variant across human populations (**Figure 3A**). We found that while the frequency of the derived “G” allele is less than 15% in African populations, it is ∼50% in Eurasian populations. This geographical distribution suggests that the variant arose before the out-of-Africa dispersal (estimated to be at ∼85 kya; [32]) but increased in frequency during the Eurasian expansion of modern humans.

To visualize the haplotypic diversity in the locus containing rs2242416, we constructed a haplotype network using SNVs surrounding rs2242416 in multiple populations, including Eurasian populations where the allele frequency is highest (**Figure 3B**). The network revealed two distinct haplotype groups, one carrying the derived “G” allele and the other carrying the ancestral “A” allele, with the derived group showing markedly reduced variation, consistent with a recent selective sweep.

To assess whether the haplotypic diversity shown in **Figure 3B** is unexpected under drift, we calculated the integrated haplotype score (iHS) for rs2242416 in multiple populations [33]. For each population, we compared the iHS values for rs2242416 against the distribution of iHS values calculated for frequency-matched polymorphisms across chromosome 6 (the chromosome on which *CRIP3* resides). We find that rs2242416 exhibits a higher-than-expected iHS value in most European populations (p < 0.0001), but not in other populations **(Figure 3C)**. This result is consistent with the hypothesis that *CRIP3* has evolved under positive selection in populations.

To further evaluate whether the elevated allele frequency in Europeans reflects recent selection, we analyzed 9,990 ancient genomes spanning four temporal bins (T4: 40–50 kya, T3: 30–40 kya, T1: 10–20 kya, T0: 0–10 kya; see Methods). The derived “G” allele increased from ∼7% at T4 to ∼53% by T3, remaining consistently high thereafter, suggesting that the putative selective sweep occurred between 50 and 30 kya. To test whether this trajectory is unusual, we compared it to 1,000 random SNVs with similar present-day frequencies (50–60%) in Europeans. Only 39 of these (3.9%) exhibited allele frequencies as low or lower than rs2242416 at T4 (**Figure 3D**). This analysis further supports the notion of positive selection acting on this locus.

The empirical outlier comparisons presented above are suggestive of selection in Europeans. However, such analyses can be biased by local mutation and recombination rate variation. Therefore, as a complementary approach, we employed CLUES [34], which uses ancestral recombination maps to infer allele-frequency trajectories over time and applies a Markov chain Monte Carlo framework to estimate the likelihood that selection explains the observed trajectory. This method is particularly well-suited for detecting strong positive selection within the last ∼5,000 years. Applying CLUES, we confirmed that the allele frequency of rs2242416 has increased steadily in Eurasian populations over the past ∼80,000 years, resulting in strong allele frequency differences between Eurasian and African populations (**Figure 3E**). However, the likelihood of selection remains small even in European populations (log-likelihood ratio = 1.9). Traditionally, values greater than 10 are interpreted as strong evidence for selection, whereas values below 0.1 reject the selection hypothesis. Accordingly, the CLUES analysis is inconclusive and strong recent selection on this locus is unlikely, at least within the last 5,000 years.

Collectively, although the allele-frequency trajectory and reduced haplotypic diversity surrounding rs2242416 are consistent with a possible selective sweep in Europeans, the CLUES results do not provide evidence for recent strong selection. Thus, further work specifically testing for weak positive selection or balancing selection (broadly defined) is warranted [35].

## Discussion

In this study, we investigated a common human haplotype harboring the non-synonymous variant rs2242416 in the *CRIP3* gene, which results in an isoleucine-to-threonine substitution at a site that is highly conserved throughout non-human mammals. We found that rs2242416 is strongly associated with age-related hearing loss and heart disease. Multiple lines of evidence support rs2242416 as the likely causal variant, including its segregation at unusually high frequency for such a conserved site and predicted alteration of hydrogen-bonding at the protein level. Population genetic analyses indicate a major rise in allele frequency, accompanied by reduced haplotypic variation at this locus in Eurasian populations during the past ∼80,000 years, although robust evidence for a recent selective sweep is lacking. Careful, simulation-based analyses will be required to test whether long-term, weak selection or balancing selection could underlie the population genetic patterns observed at this locus.

What makes rs2242416 particularly intriguing is its apparent pleiotropic influence on two functionally distinct traits: hearing and cardiovascular function. One plausible explanation is antagonistic pleiotropy, in which the variant confers early-life benefits that outweigh its detrimental effects later in life. Because age-related hearing loss typically manifests after reproduction, its fitness costs are likely minimal [36].In contrast, potential early-life benefits on cardiovascular performance could have been subject to positive selection (**Figure 4**). Specifically, the derived “G” allele, predicted to introduce a stabilizing hydrogen bond into the CRIP3 protein, may enhance cardiovascular or metabolic function. This model parallels well-known cases in which alleles that increase risk for late-onset disorders, such as Alzheimer’s disease, persist because they provide advantageous effects earlier in life [37-39]. Consistent with this idea, rs2242416 is also associated with anthropometric and developmental traits, including body-mass index and height, suggesting possible developmental-stage or tissue-specific effects. Interestingly, three lemur species and two lagomorph species independently harbor the same non-synonymous change, raising the possibility of convergent evolution at this site, potentially reflecting similar selective pressures across mammalian lineages.

**Figure 4.**
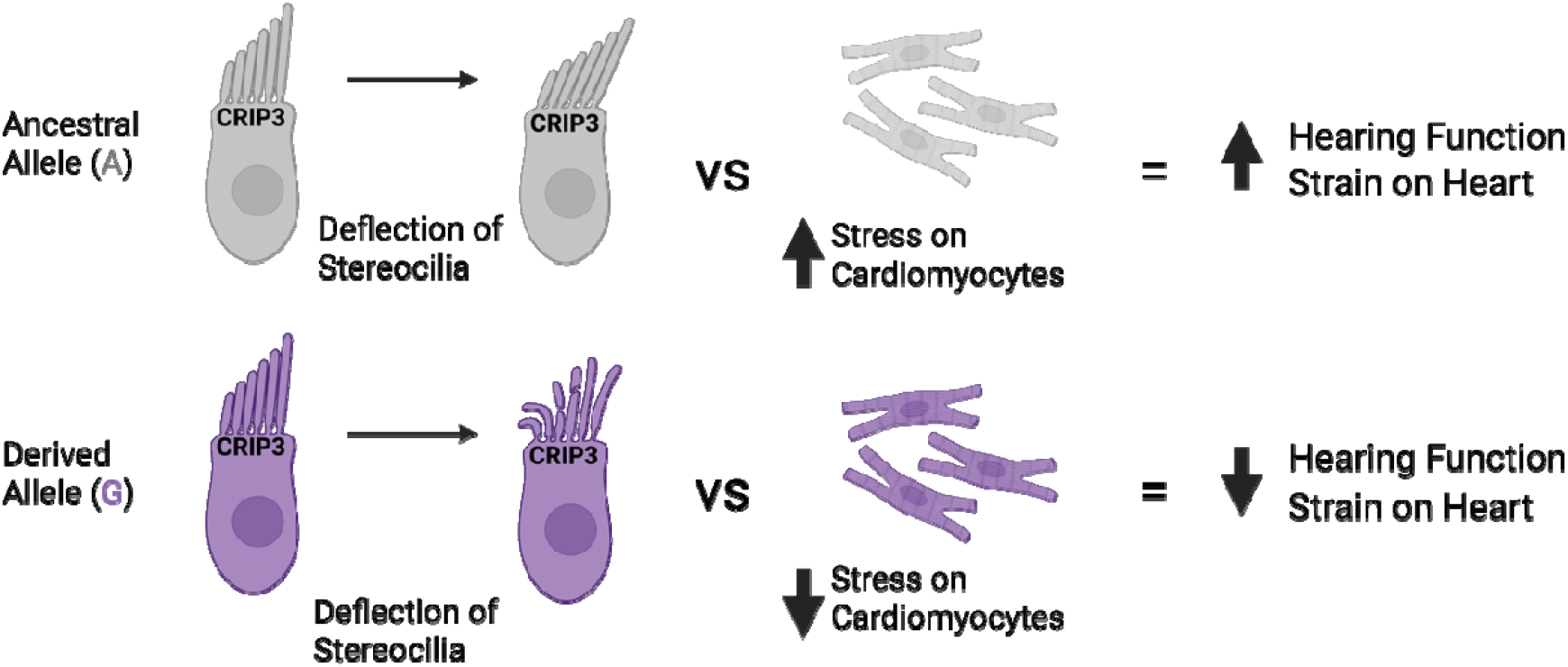
Proposed functional framework based on allele distribution. Top: Under conditions with the ancestral allele A, hearing is intact but there may be an increase strain on heart function. Bottom: Under conditions with the derived G allele (rs2242416), hearing function is compromised but there may be less stress on heart function. Figure 4 was created in BioRender. Brongo, S. (2026) https://BioRender.com/izx5bm9.

An alternative explanation is that the rise in frequency of the derived “G” allele may reflect relaxed selection on this site: the ancestral “A” allele may have been maintained by past selective pressures that weakened or disappeared within the last ∼80,000 years, allowing the derived allele to drift upward in frequency without conferring a fitness advantage. Another possibility is a reduction in the effective population size in Eurasians following the out-of-Africa migration that made purifying selection inefficient. Regardless of any direct fitness effects, the pleiotropic impact of this variant on human biology and health remains important. Future work using cellular or animal models will be needed to test these hypotheses.

Overall, our results cautiously support antagonistic pleiotropy as the most parsimonious explanation for the evolution of rs2242416, aligning with prior evidence that variants affecting cardiovascular function may have been evolving in an antagonistic pleiotropy framework at the intersection of health and evolutionary fitness [40, 41]

## Conclusion

This study provides compelling evidence that rs2242416, a non-synonymous variant in *CRIP3*, rose in frequency through a selective sweep in European populations and contributes to age-related hearing loss and heart disease. Identifying variants within their evolutionary context can help pinpoint loci with true physiological impact. Such variants may refine population-specific disease-risk models and guide experimental validation through CRISPR editing or knock-in animal models. In this context, rs2242416 serves not only as a genetic risk factor for hearing loss and heart disease but also as a model for understanding the evolutionary trade-offs embedded in the human genome. Integrating evolutionary analysis with biomedical research will continue to illuminate how adaptive history influences present-day disease risk, and how such insights can inform future therapeutic strategies.

## Methods

GWAS ATLAS and published literature was used to find our gene of interest. The search criteria involved Domain: Ear, Nose, Throat, and Trait: Hearing difficulties/problems [42]. For our interest, we looked at genes with high conservation across lineages and expression in mice, in hopes for future use in functional studies. *CRIP3* was identified as being associated with age-related hearing loss and hearing impairment and interestingly, has a SNV, rs2242416, also associated with hearing loss [7-9]. FinnGen r12 was used to validate the association of CRIP3 and rs2242416 with hearing loss [25]. A haplotype block (chr6: 43,273,604 ± 50 kilobases (kb)) from the hg19 reference genome containing *CRIP3* and rs2242416 and population data from the 1000 Genomes Project was used to calculate linkage disequilibrium values [43]. Combined Annotation Dependent Depletion (CADD) scores for the top 10 variants in strong linkage disequilibrium (R2 > 0.9) with rs2242416 were calculated using the CADD website through the University of Washington [27].

The GnomAD database and 1000 Genomes Project was used to verify allele frequencies in human populations [44]. The UCSC genome browser was used to visualize both *CRIP3* and rs2242416, and tracks including conservation data was used, specifically CACTUS align, base wise conservation, and conservation of 100 vertebrates [45]. Tracks were downloaded and WebLogo 3 was used to visualize the conserved sequences [46]

Data from the Allen Ancient DNA Resource was obtained to understand the evolutionary history of rs2242416 [47]. Version v54.1.p1 was used, which contained 16389 unique individuals, with 9990 ancient and 6399 present-day individuals [47].Based on how many individuals were available, time points were generated. T0 (0-10,000 years before present) includes 10,600 individuals, T1 (10-20,000 years before present) includes 100 individuals, T2 (20-30,000 years before present) was not included due to lack of individuals, T3(30-40,000 years before present) includes 38 individuals, T4 (40-50,000 years before present) includes 16 individuals.

For selection analysis, we calculated the integrated haplotype score (iHS) for various polymorphisms on chromosome 6, including rs2242416, in 12 populations using selscan [48]. For each population, we computed iHS values for variants on chromosome 6 that fall within the same derived allele frequency bin as rs2242416 in that population. We then standardized the iHS values in each population by calculating the Z-scores (standardized iHS) across the polymorphisms used for that population. To test the hypothesis that the derived allele (G) is under positive selection, we generated density plots of standardized iHS values for each population. If the Z score for rs2242416 is greater than 1.959964 (corresponding to a cumulative probability of 97.5%), the derived allele was interpreted to have evolved under positive selection.

Alphafold3 was used to predict and visualize the structure of the CRIP3 protein, with both the ancestral isoleucine and the derived threonine created by the rs2242416 variant [28]. Expression data from GTEx was used to understand gene expression of *CRIP3* and impacts of rs2242416 expression for specific tissues [21].

## Data Availability

No new data is produced.

## Data availability

We obtained the phenotypic association data from GWAS ATLAS (https://atlas.ctglab.nl/) and FinnGen (https://r12.finngen.fi/variant/6:43305866-A-G). CADD scores were obtained from (https://cadd.gs.washington.edu/). Population genetics analysis was performed using the 1000 genomes data obtained from (https://www.internationalgenome.org/category/data-access/). Bulk RNA-seq expression data (TPM, GTEx v8) is available from the GTEx open-access portal (https://gtexportal.org/home/downloads/adult-gtex/bulk_tissue_expression). No new data was produced in this study.

## Acknowledgements

O.G. acknowledges support from the National Science Foundation (under grant nos. 2049947 and 2123284) and the National Institute of Health (R35-GM156519). The funders had no role in study design, data collection and analysis, publication decision, or manuscript preparation.

## Notes

### Competing Interest Statement

The authors have declared no competing interest.

### Funding Statement

NSF 2049947 & 2123284
National Institute of Health (R35-GM156519)

### Author Declarations

1000 Genomes Project GWAS Atlas GTEx consortium

