## Supplementary tables and figures for "Genetic and functional evidence implicates a *CRIP3* non-synonymous variant in age-related hearing loss"

**Table S1. Top 10 PheWAS hits associated with rs2242416.** The top phenotypic association with rs2242416 is “hearing difficulty/problems”. The effect allele for this trait is the “G” allele. Other top associations include cardiovascular traits, with the effect allele being the “A” allele.

| **atlas ID** | **PMID** | **Year** | **Domain** | **Trait** | **p-value** | **N** | **EA** | **NEA** |
| --- | --- | --- | --- | --- | --- | --- | --- | --- |
| **3313** | 31427789 | 2019 | Ear, Nose, Throat | Hearing difficulty/problems | 3.69E-18 | 370713 | G | A |
| **4043** | 30124842 | 2018 | Skeletal | Height | 5.50E-18 | 693529 | G | A |
| **4077** | 30239722 | 2018 | Metabolic | Waist-hip ratio | 3.56E-17 | 697734 | A | G |
| **3314** | 31427789 | 2019 | Ear, Nose, Throat | Hearing difficulty/problems with background noise | 3.35E-16 | 378722 | G | A |
| **4080** | 30239722 | 2018 | Metabolic | Waist-hip ratio (adjusted for BMI) | 9.92E-14 | 694649 | A | G |
| **3187** | 31427789 | 2019 | Skeletal | Standing height | 3.78E-13 | 385748 | G | A |
| **3551** | 31427789 | 2019 | Cardiovascular | Vascular/heart problems diagnosed by doctor: High blood pressure | 2.18E-12 | 385699 | A | G |
| **4142** | 29403010 | 2018 | Cardiovascular | Systolic Blood Pressure | 2.97E-12 | 136597 | A | G |
| **4079** | 30239722 | 2018 | Metabolic | Waist-hip ratio (female) | 2.06E-11 | 381152 | A | G |
| **4144** | 29403010 | 2018 | Cardiovascular | Mean arterial pressure | 3.98E-11 | 136482 | A | G |

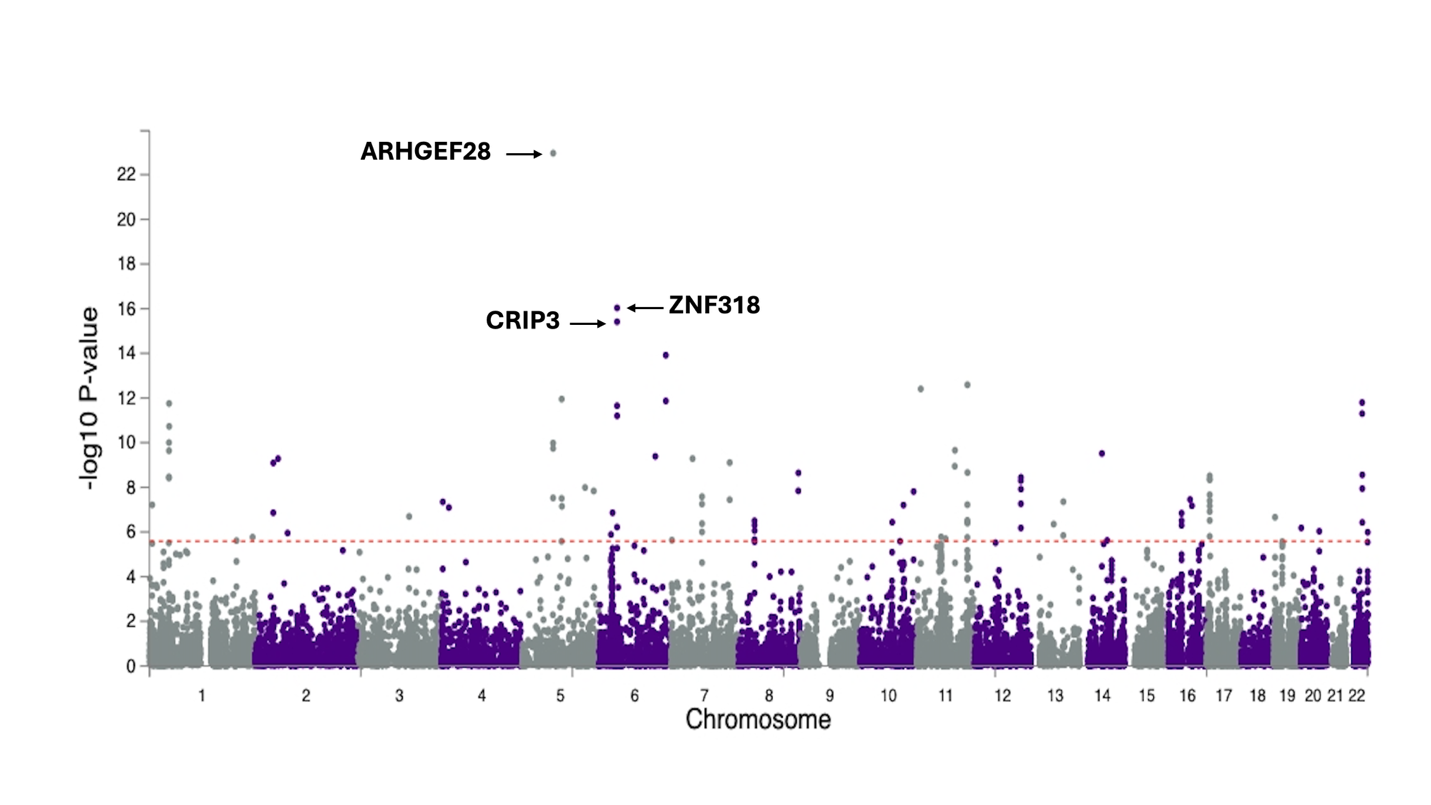

**Figure S1. Top GWAS hits for hearing loss.** Manhattan plot shows the top 3 genes associated with hearing loss. Red line indicates significance and the top 3 genes are labeled
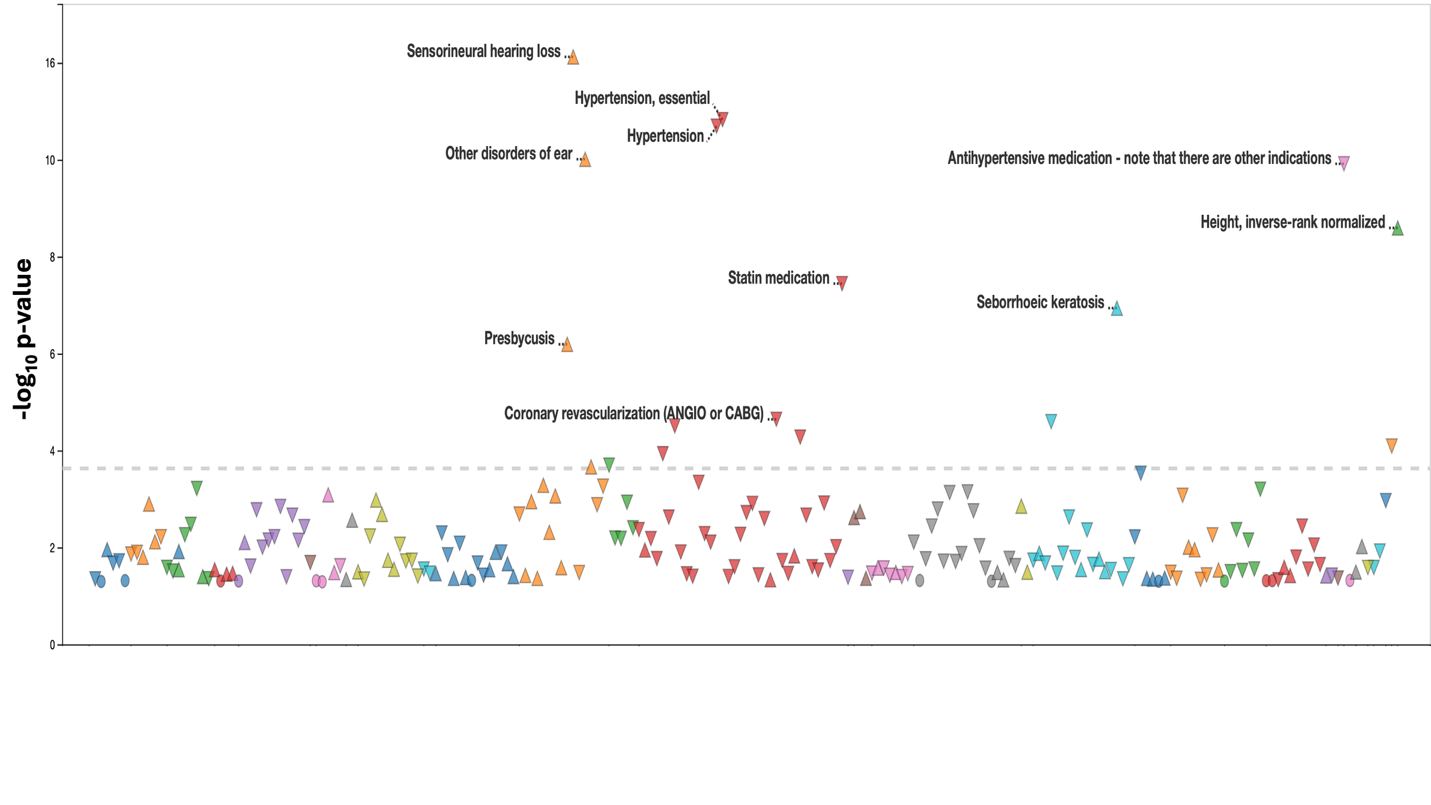

**Figure S2. Confirmation of association in FinnGen Database.** Association of phenotypic traits with rs2242416. Notably, the most association trait is hearing loss.

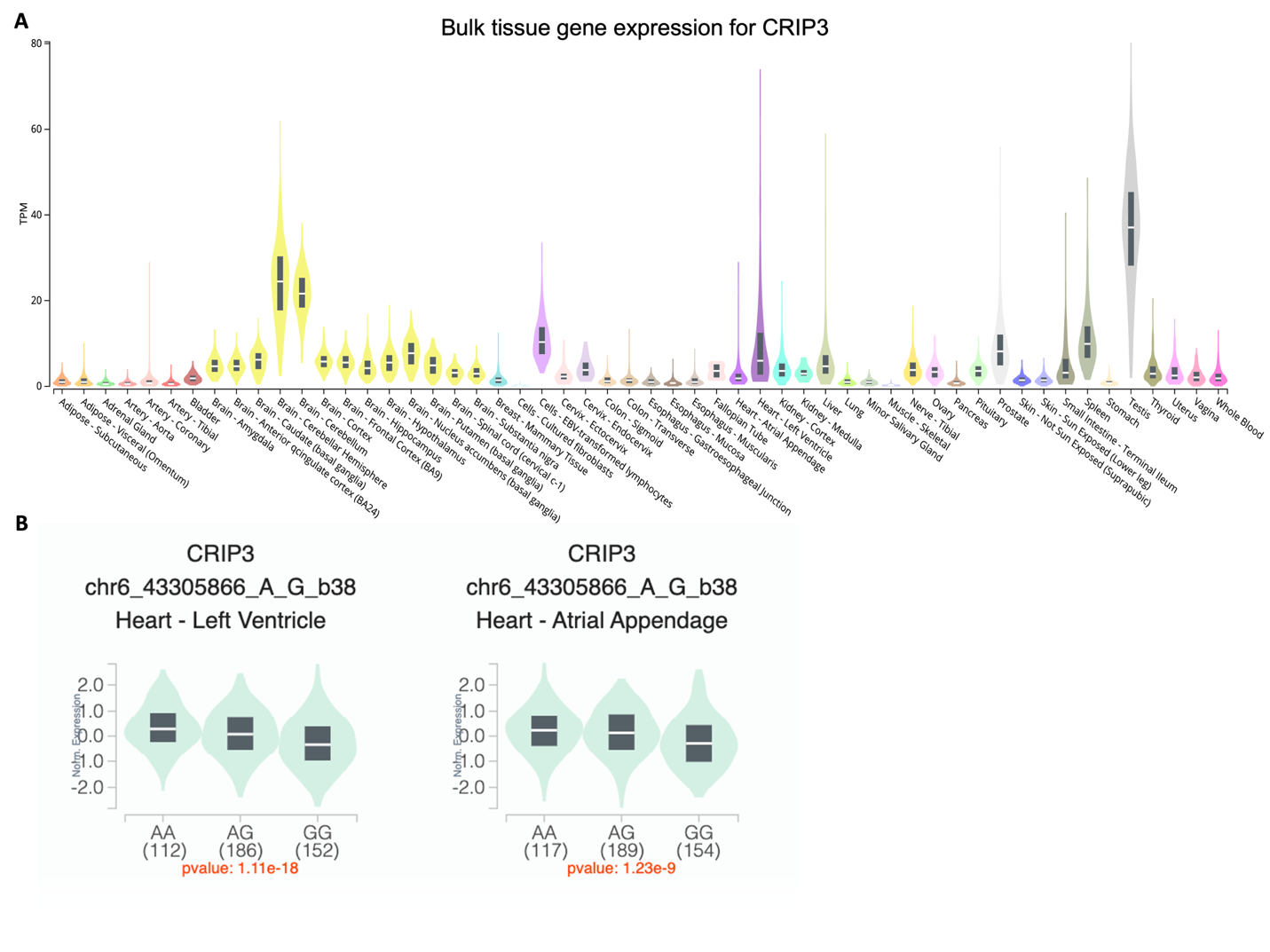

**Figure S3. Expression Data of CRIP3 in multiple tissues and consequences of rs2242416 on expression.** A. CRIP3 is widely expressed with notable areas of the brain, heart, and spleen. B. Expression Quantitative Trait Loci indicate a decrease in expression of CRIP3 in the left ventricle when rs2242416 is present.

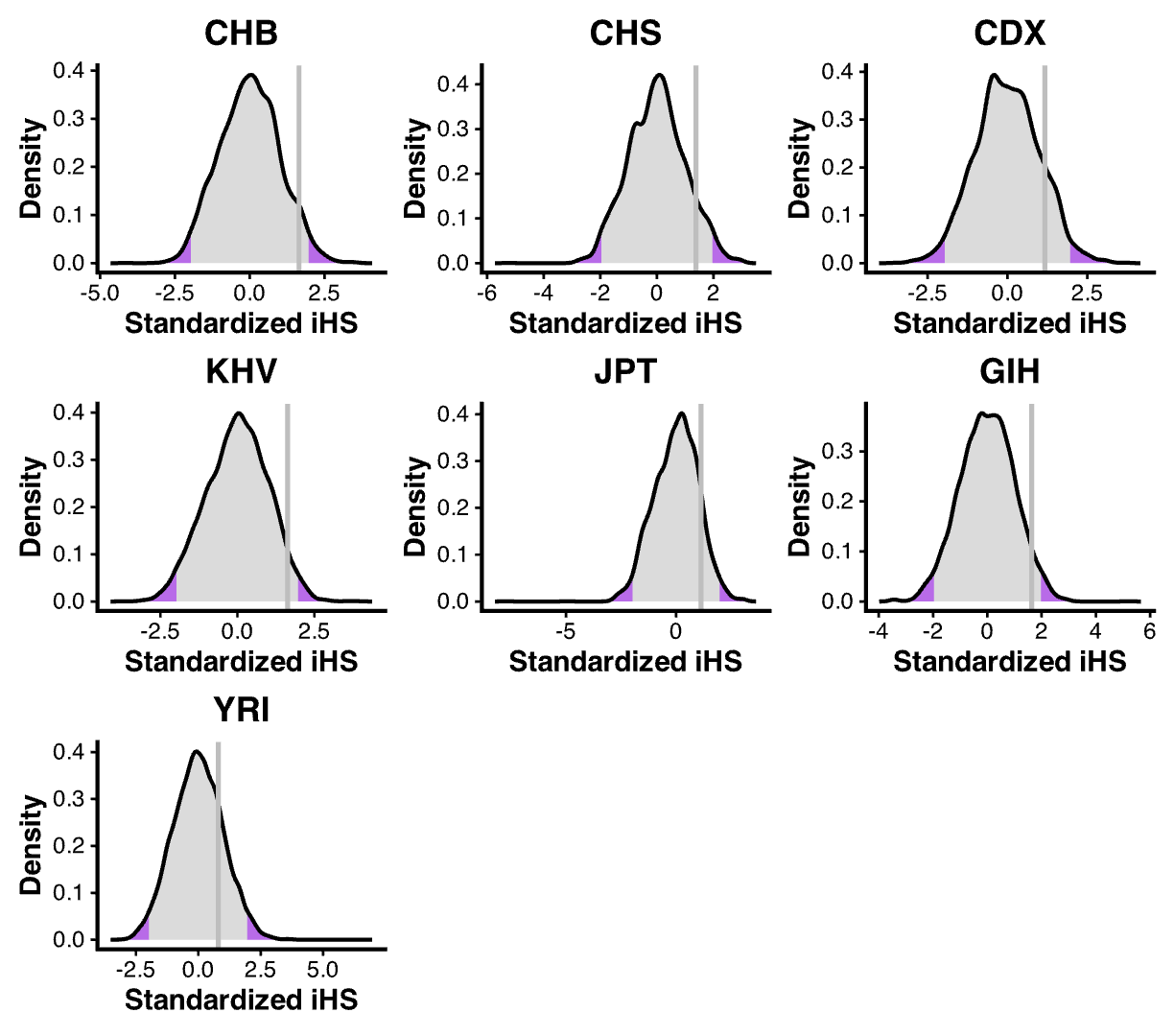

**Figure S4. IHS Values for representative populations.** Neither East Asian, South Asian, or African populations detected a selection signal. Population codes: CHB: Han Chinese in Beijing, China, CHS: Han Chinese South, CDX: Chinese Dai in Xishuangbanna, China, KHV: Kinh in Ho Chi Minh City, Vietnam, JPT: Japanese in Tokyo, Japan, GIH: Gujarati Indian in Houston, Texas, YRI: Yoruba in Ibadan, Nigeria.
